# Area deprivation, social vulnerability, and post-pregnancy blood pressure

**DOI:** 10.64898/2026.02.10.26346041

**Authors:** Allison E. Gaffey, Matthew M. Burg, Andrea C. Kozai, Virginia R. Nuckols, Jun Wu, Kiarri Kershaw, William Grobman, Bethany Barone Gibbs, the National Heart, Lung, and Blood Institute nuMoM2b Health Heath Study Network

## Abstract

**Introduction:** Pregnancy is a critical test of women’s cardiovascular risk. Structural factors may influence long-term cardiovascular health beyond individual, social experiences. We examined associations of neighborhood-level deprivation and individual-level social vulnerability (SV) during pregnancy with postpartum blood pressure (BP).

**Methods:** This secondary analysis of a prospective cohort study used data from 3,728 nulliparous women in the nuMoM2b-HHS cohort followed from early pregnancy to 2-7 years post-delivery (Mage: 30.8 years, 65% non-Hispanic White, 14% with adverse pregnancy outcomes [APOs]). Multivariable linear and logistic regression models tested relations of the Area Deprivation Index (ADI) and SV (a composite of perceived stress, discrimination, pregnancy experiences, social support, health literacy, depression, and anxiety) with systolic BP (SBP), diastolic BP (DBP), and incident hypertension, adjusting for demographic and behavioral covariates. Effect modification by APO history was assessed.

**Results:** In unadjusted models, both ADI and greater SV were positively associated with SBP and DBP (all *ps*<0.001). After adjustment, ADI remained positively associated with BP: each 10-unit increase in ADI was associated with 1.0 mmHg higher SBP (*p*=0.008) and 0.6 mmHg higher DBP (*p*=0.013). However, SV was no longer associated with BP after adjustment. ADI and SV were not associated with incident hypertension. No evidence of effect modification by APO history was observed (interactions *p*>0.20).

**Conclusions:** Neighborhood deprivation during pregnancy was associated with higher BP up to seven years later, independent of individual social vulnerability. Structural context during pregnancy may contribute to early maternal cardiovascular risk.

## Introduction

Pregnancy offers a unique “window to future health,” revealing early biological and clinical indicators of women’s long-term cardiovascular risk.^1^ Both the American Heart Association (AHA) and the American College of Cardiology (ACC) now emphasize pregnancy history as a vital component of cardiovascular disease (CVD) risk assessment in women.^2,4^ For instance, adverse pregnancy outcomes (APOs), such as hypertensive disorders of pregnancy, gestational diabetes, preterm delivery, and fetal growth restriction, are well-established predictors of later CVD.^2^ Yet, APOs do not fully capture the broader environmental and social contexts that shape women’s cardiovascular trajectories across the life course.^23^

Structural determinants of health, particularly neighborhood socioeconomic disadvantage or deprivation, are increasingly recognized as powerful and durable contributors to cardiovascular risk, and extend beyond the perinatal period.^5^ Neighborhood deprivation, often quantified using indices such as the Area Deprivation Index (ADI), is a composite of income, education, employment, and housing quality that captures these community-level exposures.^6^ Recent validation work demonstrates that ADI and related neighborhood risk indices robustly predict morbidity and mortality across diverse populations, underscoring their utility as clinically meaningful measures of structural risk.^7^ Women who reside in highly disadvantaged neighborhoods during pregnancy face significantly greater 30-year CVD risk and a greater burden of subclinical atherosclerosis than those in more advantaged areas, independent of individual health behaviors.^8,9^ This structural risk may be incurred by limited access to healthy foods, reduced opportunities for safe physical activity, and heightened, chronic exposure to environmental and interpersonal stressors.^5^

In parallel, individual-level social vulnerability (SV), which encompasses cumulative adverse social exposures such as discrimination and psychosocial stress is consistently associated with elevated blood pressure (BP) and a greater risk for hypertension among women. ^10–14,15^ SV closely intersects with the broader social determinants of health, which encompass one’s economic stability, educational attainment, neighborhood context, and social environment.^16^ Notably, SV may be disproportionately prevalent among women from racially and socioeconomically marginalized groups, contributing to persistent disparities in maternal and cardiovascular outcomes.^15,17^ However, the relative contributions of neighborhood-level structural disadvantage versus individual-level social vulnerability during pregnancy to postpartum BP remain poorly understood. Furthermore, APOs are increasingly conceptualized as markers of underlying vascular and metabolic susceptibility, which may heighten sensitivity to the effects of adverse environmental and social exposures in the years following pregnancy. The primary objective of this study was to evaluate the association of both neighborhood-level deprivation and individual-level SV during pregnancy to BP and risk for hypertension measured 2 to 7 years after delivery. We hypothesized that neighborhood deprivation and individual SV in pregnancy would each be independently associated with elevated BP in the years following pregnancy, even after adjusting for conventional confounders. As a secondary objective, we evaluated if these associations differed by history of APOs, to assess whether structural and social risk factors exert differential associations with post-pregnancy BP among women with prior pregnancy complications.

## Methods

### Study Population

The present analyses were conducted using data from the Nulliparous Pregnancy Outcomes Study-Monitoring Mothers-to-Be Heart Health Study (nuMoM2b-HHS).^18^ This was a longitudinal cohort study in which participants were recruited during the first trimester of their first pregnancy and followed into the postpartum period. The cohort was recruited across 8 U.S. clinical centers (2010-2013 for nuMoM2b, 2014-2020 for HHS). Women enrolled were assessed at multiple time points across the perinatal period, with a follow-up visit conducted 2 to 7 years after delivery. Women were excluded from the current analysis if they had hypertension or CVD at baseline, if their pregnancy ended prior to 20 weeks’ gestation due to perinatal death, or if they were missing follow-up data or covariates (Figure 1). For the secondary aim, women were excluded if they were missing information concerning APOs. Thus, the analytic sample included women with complete BP data from the first trimester of pregnancy, and the post-pregnancy visit and available measures for SV, ADI, and relevant covariates. The sample size was determined by the number of eligible participants in the nuMoM2b-HHS cohort with complete exposure, outcome, and covariate data; no *a priori* power calculation was performed for this secondary analysis.

**Figure 1.**
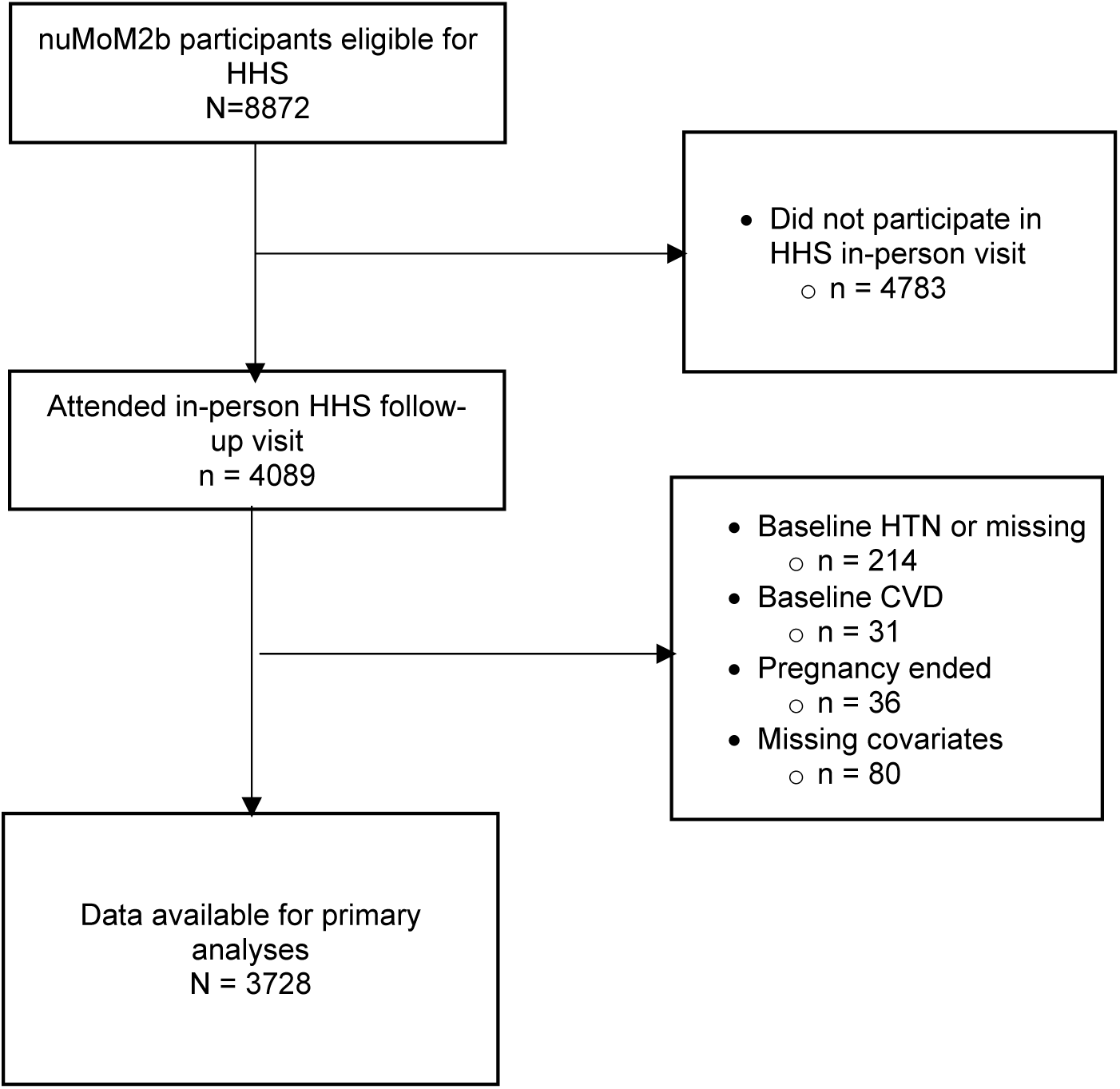
Flow diagram of sample retention for analyses. Flowchart depicting the derivation of the analytic sample. Starting from the full nuMoM2b enrollment cohort, women were sequentially excluded if they did not participate in the Heart Health Study follow-up visit or had missing blood pressure or predictor data. The final analytic cohort consisted of 3,728 women, of whom 541 experienced an adverse pregnancy outcome (APO) and 3,187 did not.

### Measures

#### Neighborhood Socioeconomic Disadvantage - Area Deprivation Index (ADI)

The primary predictor of interest was neighborhood socioeconomic disadvantage. This factor was assessed using the ADI, which is a validated, composite measure of neighborhood-level socioeconomic disadvantage that is derived from U.S. Census data.^19^ The ADI incorporates 17 variables reflecting income, education, employment, and housing quality. The variables are aggregated at the Census block-group level and standardized to national percentiles, with higher percentiles indicating greater deprivation. For this analysis, participants’ residential addresses at enrollment in the nuMoM2b cohort during the first trimester of pregnancy were geocoded and linked to the corresponding ADI value using the Neighborhood Atlas (University of Wisconsin School of Medicine and Public Health).^19^ As in prior nuMoM2b analyses, ADI was examined continuously, and also categorized into quartiles for analytic comparisons, with the highest quartile representing the most disadvantaged neighborhoods.^20^

#### Individual-level Social Vulnerability (SV)

The second predictor of interest was a study-specific composite measure of SV during pregnancy, constructed *a priori* using seven domains selected to capture multidimensional, individual-level social exposures available in the nuMoM2b cohort. These domains were chosen based on prior literature and data availability. SV components were measured during the first trimester of pregnancy, unless otherwise stated. Domains included: (1) perceived stress, measured with the Cohen 10-item Perceived Stress Scale;^21^ (2) discrimination, assessed with the Experiences of Discrimination questionnaire;^22^ (3) pregnancy experiences, assessed using the Difficulties in Pregnancy subscale of the Pregnancy Experience Scale;^23^ (4) social support, assessed with the Multidimensional Scale of Perceived Social Support;^24^ (5) health literacy, assessed using the Rapid Assessment of Adult Literacy;^25^ (6) depression, assessed with the Edinburgh Postnatal Depression Scale;^26^ (7) anxiety assessed with the State-Trait Anxiety Inventory – Trait Subscale.^27^ For each domain, factor scores were derived and standardized to z-scores. These domain-specific z-scores were then combined to generate a latent SV construct, which was used to reduce measurement error and limit multiple testing.

#### Blood Pressure Outcomes

The primary outcomes were SBP and DBP measured at the 2- to 7-year post-delivery visit.^28^ The secondary outcome was incident postpartum hypertension defined as Stage 1 or higher hypertension (SBP ≥130 mm Hg or DBP ≥80 mm Hg) or Stage 2 hypertension (SBP ≥140 mm Hg or DBP ≥90 mm Hg) according to the 2025 clinical guidelines,^29^ or self-reported use of antihypertensive medication during the postpartum follow-up period. BP was assessed using a standardized research protocol.^18^ Trained study staff obtained three seated measurements after a 5-minute rest period using calibrated automated oscillometric monitors (OMRON HEM-907XL, Omron Healthcare Incorporated, Lake Forest, Illinois). The mean of the final two SBP and DBP readings was used for analysis.

#### Covariates

All models were adjusted for maternal age at delivery, race/ethnicity, body mass index (BMI) and cigarette smoking measured at the nuMoM2b-HHS baseline visit, first-trimester systolic BP, and hypertensive disorders of pregnancy.

#### Bias

Several strategies were used to reduce potential bias. BP was measured using a standardized protocol by trained staff using calibrated devices to minimize measurement error. Prospective collection of exposure data during early pregnancy reduced recall bias. Multivariable models adjusted for established demographic and clinical confounders, including baseline SBP, to mitigate confounding. Still, residual confounding from unmeasured variables was possible.

### Statistical Analysis

All statistical tests were two-sided, with α set to 0.05. Analyses were conducted using SAS Version 7.4 (SAS Institute, Cary, North Carolina). Participants with missing exposure, outcome, or covariate data were excluded from analyses (complete-case analysis). The proportion of missing data was low for most variables. Descriptive statistics were calculated for baseline characteristics and presented both for the total sample and stratified based on APO history. Continuous variables were summarized as means ± standard deviations (SD) and categorical variables were summarized as frequencies with percentages.

All multivariable analyses were first conducted in the full sample. To assess the associations between pregnancy SV and BP outcomes several years after delivery, multivariable linear regression was used for continuous BP measures, and multivariable logistic regression was used for binary hypertension outcomes. The primary analyses were modeled in three ways, with either SBP or DBP as the outcome of each. First, models included the SV factor as the primary predictor. Second, models included ADI as the primary predictor. And third, models were conducted including both SV and ADI simultaneously. To improve interpretability, ADI was scaled per 10-unit increase in all regression models. Secondary analyses included incident hypertension as the outcome variable. Exploratory analyses were used to evaluate whether associations differed by APO history, if which we tested effect modification by including interaction terms between APO status and each exposure (ADI and SV measures) in covariate-adjusted models. Stratified estimates by APO status were considered descriptive and are presented only as supplemental material.

Finally, sensitivity analyses were used to examine associations of individual SV components with BP, using a Bonferroni correction. Models were also conducted while excluding women who experienced an intervening pregnancy between the index delivery and the nuMoM2b-HHS follow-up visit to minimize the potential influence of subsequent reproductive events. Additional subgroup comparisons were performed between women without an APO in the index pregnancy and two mutually exclusive groups: (1) women with a hypertensive disorder of pregnancy, (2) women with any non-hypertensive APO.

## Results

### Baseline Characteristics

Among 3,728 participants in the nuMoM2b-HHS cohort, the average age was 30.8 years, and 70% of the cohort were in the 22-35 age group (Table 1). Overall, 65% of the women were non-Hispanic White and 16% were Hispanic. Average follow-up from delivery to the HHS visit was 3.2 years. Across the cohort, the average BMI was 26, with 57% of participants in the 18-25 category (i.e., normative). Early in pregnancy, the average BP was 109/67 mm Hg. About 14% of women experienced an APO during their nuMom2b pregnancy, and those women differed from without such a history on most factors. Sociodemographic differences were present such that those with APOs were more likely to belong to racial and ethnic minority groups, be unmarried, reported lower educational attainment, and more often relied on government insurance, compared to those who did not experience an APO. Women with an APO also exhibited greater SBP (112.0 vs 108.4 mmHg, *p* < .001) and DBP (69.6 vs 66.6 mmHg, *p* < .001), and less favorable other cardiovascular risk factors. Missing data were minimal for primary exposure and outcome variables (<2% for all key variables).

**Table 1.**
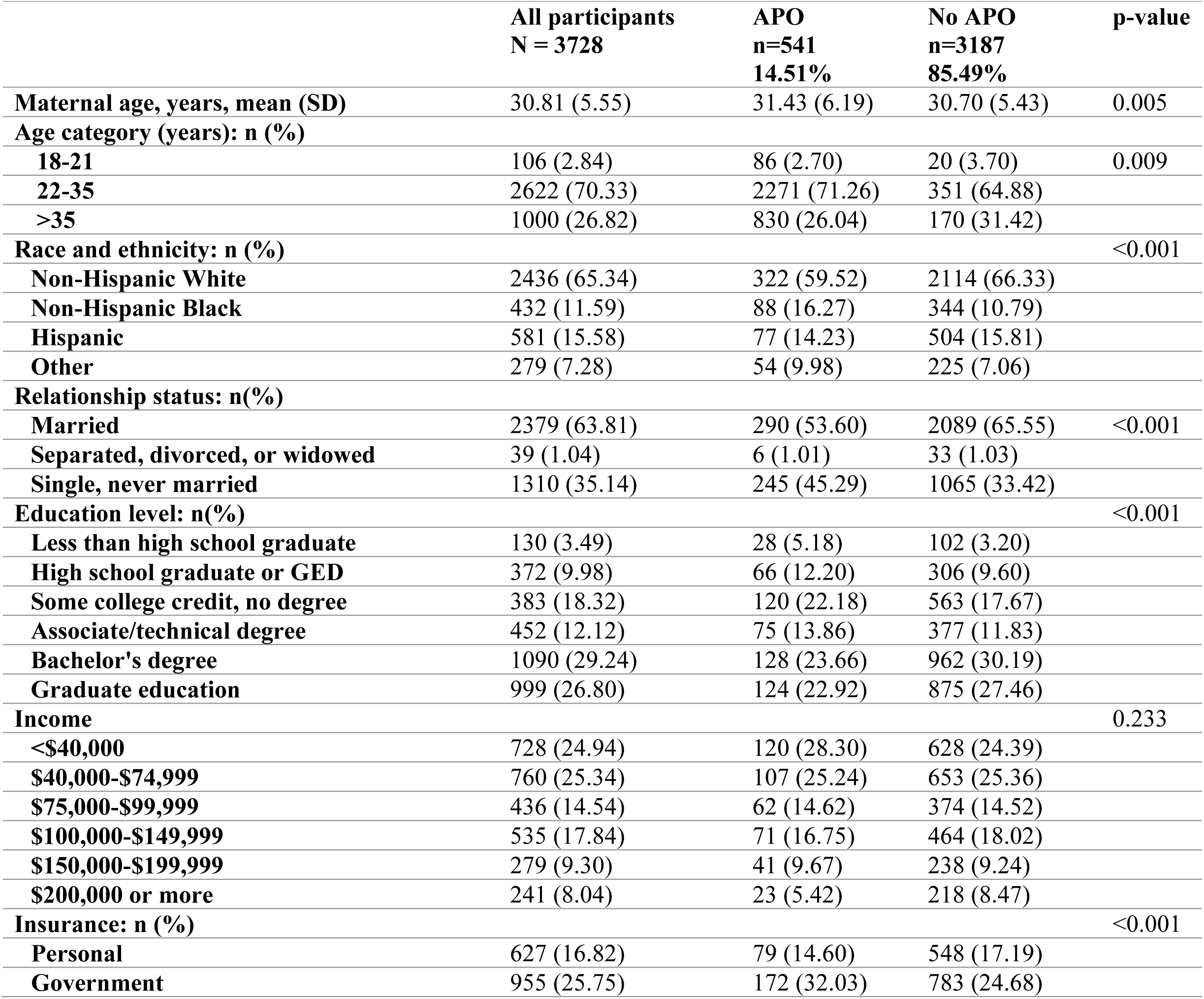
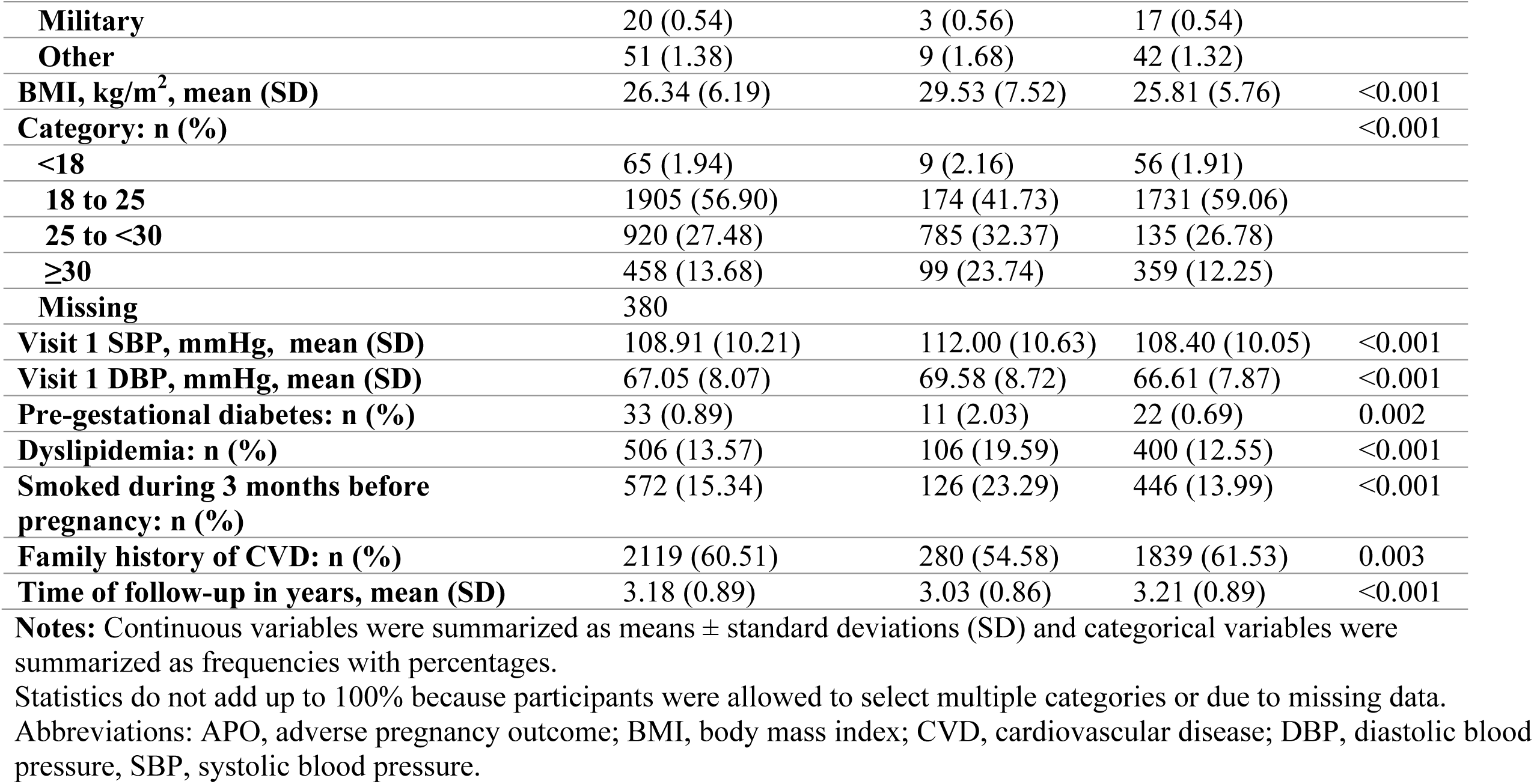
Baseline demographic and pregnancy characteristics of the nuMoM2b-HHS sample.

Baseline characteristics by ADI and SV quartiles are depicted elsewhere (Supplemental Tables 1 and 2). SBP and DBP were progressively higher across quartiles of neighborhood deprivation defined by ADI. Participants in the highest ADI quartile had modestly higher mean SBP/DBP values (112.7/72.9 mmHg) compared with those in the lowest quartile (110.6/71.1 mmHg; ps<0.001). In contrast, BP values varied minimally across SV quartiles, with only small differences in mean SBP (111.3-111.6 mmHg) and DBP (71.5-72.6 mmHg), despite statistically significant associations (*p* < 0.001) which were likely due to the large sample size.

### SV Component Description

The SV index developed for this study demonstrated strong and consistent gradients for each component across quartiles (Supplemental Table 3). Women in higher SV quartiles reported substantially greater psychosocial stressors, including perceived stress (5.1 in Q1 vs 16.8 in Q4; *p* < 0.001), discrimination (0.21 vs 0.60; *p* < 0.001), pregnancy-related difficulties (0.49 vs 0.79; *p* < 0.001), postpartum depression (2.1 vs 9.0; p < 0.001), and anxiety (25.6 vs 42.5; p < 0.001). Conversely, social support and health literacy was lower across quartiles (social support: 82.5 in Q1 vs 61.5 in Q4, *p* < 0.001; health literacy: 6.9 vs 6.5, *p* < 0.001).

### Associations of ADI and SV With Postpartum BP

Next, multivariable models were used to examine ADI and SV continuously in association with BP. In separate, unadjusted models of ADI and SV, both variables were again positively associated with SBP and DBP at follow-up (ps<0.001; Table 2). After models were adjusted for demographic and clinical covariates, each 10-unit increase in the continuous ADI score was associated with higher SBP (*β* = 0.54, *95% CI*: 0.18,0. 89, *p* = 0.002) and DBP (*β* = 0.41, *95% CI*: 0.10,0.72, *p* = 0.008). In contrast, SV was no longer significantly associated with BP (SBP: *β* = -0.22, *95% CI:* -3.57,3.12, *p* = 0.893; DBP: *β* = 0.33, *95% CI:* -2.60,3.27, *p* = 0.824). When ADI and SV were modeled jointly in association with BP, only ADI remained a significant predictor of follow-up SBP (*p* = 0.001) and DBP (*p* = 0.006). These associations were robust when ADI was modeled both as a continuous measure and across quartiles of neighborhood deprivation.

**Table 2.** Associations of SV and ADI with blood pressure at 2-7 years postpartum.

| SBP |  |  |  | DBP |  |  |
| --- | --- | --- | --- | --- | --- | --- |
| Separate ADI and SV models |  |  |  |  |  |  |
| | $\beta$(SE) | 95% CI | p | $\beta$(SE) | 95% CI | p |
| ADI | 0.97 (0.20) | 0.57,1.36 | <0.001 | 0.58 (0.18) | 0.24,0.93 | 0.001 |
| SV | -0.40 (0.04) | -0.78,0.04 | 0.029 | -0.32 (0.17) | -0.64,0.01 | 0.054 |
| ADI x APO | 0.09 (0.18) | -0.24,0.42 | 0.584 | -0.04 (0.15) | -0.33, 0.26 | 0.810 |
| SV x APO | -0.70 (0.50) | -1.68,0.28 | 0.162 | 0.19 (0.44) | -0.68,1.06 | 0.671 |
| SBP |  |  |  | DBP |  |  |
| ADI and SV models |  |  |  |  |  |  |
| | $\beta$(SE) | 95% CI | p | $\beta$(SE) | 95% CI | p |
| ADI | 0.94 (0.17) | 0.54, 1.33 | <0.001 | 0.56 (0.18) | 0.01, 0.08 | <0.001 |
| SV | -0.35 (0.19) | -0.73, 0.02 | 0.064 | -0.27 (0.17) | -0.60, 0.06 | 0.107 |
| ADI x APO | 0.09 (0.17) | -0.24, 0.42 | 0.584 | -0.04 (0.15) | -0.33, 0.26 | 0.810 |
| SV x APO | -0.63 (0.50) | -1.61, 0.35 | 0.208 | 0.20 (0.45) | -0.68, 1.07 | 0.647 |
| ADI x SV x APO | 0.29 (0.13) | 0.03, 0.55 | 0.031 | 0.27 (0.12) | 0.04, 0.50 | 0.019 |

**Notes:**
P-values are based on covariate-adjusted linear mixed models of ADI, SV, or the ADI and SV composite. ADI estimates are reported per 10-unit increase. SV estimates represent a 1-SD increase in the composite score. Covariates included age, race and ethnicity, education, body mass index, baseline systolic blood pressure, and follow-up time.
Abbreviations: ADI, Area Deprivation Index; DBP, diastolic blood pressure; SBP, systolic blood pressure; SV, social vulnerability

### Associations of ADI and SV with Incident Hypertension

Next, analyses were conducted to examine the associations of ADI and SV with incident hypertension after pregnancy (Table 3). In unadjusted models, ADI was not associated with odds of developing Stage 1 (*p* = 0.265) or Stage 2 hypertension (*p* = 0.926). Similarly, SV was not related to the odds of developing Stage 1 hypertension but was significantly associated with Stage 2 hypertension (*p* = 0.039), with women in the third SV quartile demonstrating approximately twofold greater likelihood compared with those in the lowest quartile (OR = 1.79, 95% CI 0.93-3.44). After adjusting for demographic and clinical covariates, these associations were attenuated and no longer statistically significant. When ADI and SV were included in the same model, neither remained a significant predictor of incident Stage 1 or Stage 2 hypertension.

**Table 3.** Associations of ADI and SV Quartiles with Incident Hypertension at 2–7 Years Postpartum, Overall and by APO History.

| Predictor | Quartile | Stage 1 HTN |  | Stage 2 HTN |  |
| --- | --- | --- | --- | --- | --- |
|  |  |  | *p* |  | *p* |
| ADI | Q1 (ref) | — | — | — |  |
|  | Q2 | 0.94 (0.73–1.22) |  | 0.90 (0.51–1.59) |  |
|  | Q3 | 1.20 (0.93–1.56) |  | 1.07 (0.60–1.88) |  |
|  | Q4 | 1.07 (0.79–1.43) | 0.265 | 0.91 (0.49–1.71) |  |
| SV | Q1 (ref) | — | — | — |  |
|  | Q2 | 1.11 (0.85–1.45) |  | 1.19 (0.60–2.39) |  |
|  | Q3 | 0.97 (0.74–1.28) |  | 1.53 (0.78–2.98) |  |
|  | Q4 | 0.97 (0.73–1.28) | 0.728 | 1.79 (0.93–3.44) |  |

**Notes:** Stage 1 hypertension defined as systolic BP 130–139 mmHg or diastolic BP 80–89 mmHg. Stage 2 hypertension defined as systolic BP $\geq 140$ mmHg or diastolic BP $\geq 90$ mmHg. Models were adjusted for age, race and ethnicity, education, body mass index, baseline systolic BP, and follow-up time.
Abbreviations: ADI = Area Deprivation Index, APO = Adverse Pregnancy Outcome, CI = Confidence Interval, OR = Odds Ratio, SV = Social Vulnerability

### Exploratory Analyses based on APO History

APO history was independently associated with higher SBP and DBP (both *ps* < 0.001; Supplemental Table 4). In covariate-adjusted linear models, higher neighborhood deprivation (ADI) was associated with higher postpartum SBP (*p* = 0.008) and DBP (*p* = 0.009), whereas SV was not associated with SBP (*p* = 0.650). There was no evidence of effect modification by APO history for ADI in models of SBP or DBP (ADI×APO: *p* = 0.339 and *p* = 0.976, respectively), or for SV (SV×APO: *p* = 0.208). In adjusted logistic regression, ADI was not associated with incident hypertension (*p* = 0.516), and there was no evidence of interaction by APO history (ADI×APO: *p* = 0.730), while APO history remained strongly associated with incident hypertension (*p* < 0.001). In models of individual SV components, pregnancy-related stress showed a borderline association with incident hypertension (*p* = 0.051), but the stress×APO interaction was not significant (*p* = 0.863).

When adjusted analyses were stratified by APO history (Supplemental Table 5), there was no statistical evidence that associations differed by APO history. Expressed per 10-unit increase in ADI, associations were observed both among women with an APO history (SBP: *β* = 1.62, *95% CI*: 0.32-2.92, *p* = 0.015; DBP: *β* = 1.21, *95% CI:* 0.05-2.38, *p* = 0.042) and among women without APOs (SBP: *β* = 0.80, *95% CI:* 0.38-1.23, *p* < 0.001; DBP: *β* = 0.55, *95% CI:* 0.18-0.92, *p* = 0.004). In contrast, SV was only significantly associated with SBP in the no APO group (*β* = -0.45, *95% CI:* -0.84,-0.06, *p =* 0.023). Neither ADI or SV was associated with incident hypertension in the overall sample or within APO strata, although higher SV was associated with lower odds of incident hypertension (*OR* = 0.89, *95% CI:* 0.80–0.99, *p* = 0.035).

### Sensitivity Analyses

Next, we evaluated if individual SV components were prospectively associated with BP during follow-up (Supplemental Table 6). When all components were included simultaneously in adjusted analyses, only pregnancy-related stress was positively associated with SBP (*β* = 0.70, *95% CI*: 0.28 to 1.12, *p* = 0.001). No other SV components (i.e., perceived stress, discrimination, social support, health literacy depressive symptoms, or anxiety) were related to SBP or DBP (all *p* > 0.05).

Sensitivity analyses excluding women who experienced an intervening pregnancy between the nuMoM2b index delivery and the HHS follow-up visit yielded nearly identical results (Supplemental Table 6). In adjusted models, each 10-unit increase in ADI was associated with higher SBP (*β* = 0.28, *95% CI*: 0.13–0.44, p < 0.001) and DBP (*β* = 0.21, *95% CI*: 0.07–0.34, p = 0.002), whereas SV was not associated with either outcome (all *ps* > 0.10).

## Discussion

In this large prospective cohort of newly pregnant women who were followed up to seven years after delivery, neighborhood deprivation measured by the ADI, modeled both continuously and by quartiles, was associated with higher SBP and DBP. These associations persisted after adjustment for demographic and clinical covariates and were observed among women both with and without a history of APOs. Conversely, a composite measure of individual-level SV was not independently associated with BP or incident hypertension at follow-up. Together, these findings the importance of structural determinants,^30^ particularly structural disadvantage, in shaping long-term maternal cardiovascular health.

Neighborhood deprivation is increasingly recognized as a contributor to cardiovascular health across the life course.^30–32^ Prior work using the ADI and similar indices shows that residing in socioeconomically disadvantaged neighborhoods is associated with higher rates of hypertension, diabetes, and CVD mortality.^33–35^ Recently, a group validated eight commonly used area-based measures of social risk, including the ADI, demonstrating that neighborhood deprivation indices robustly predict both morbidity and all-cause mortality across diverse populations.^7^ Another large meta-analysis of over 250,000 UK Biobank participants (54% women) showed that those who lived in the most deprived neighborhoods also had the greatest risk of CVD incidence and related mortality.^36^ In maternal health, another recent analysis from nuMoM2b showed that women who lived in the most disadvantaged neighborhoods during pregnancy had a 10% greater 30-year CVD risk compared with those in the least disadvantaged neighborhoods.^3^ Our findings extend this literature by showing that structural disadvantage during pregnancy is associated with measurable BP elevations just a few years after a first pregnancy, suggesting that cardiovascular effects of structural context may manifest early in women’s adult life course.

In contrast with our hypothesis, a multidimensional composite of seven individual-level social and psychological components was not independently associated with postpartum BP or hypertension after adjustment. This finding differs from prior studies linking psychosocial stress, discrimination, and poor social support to cardiometabolic risk in women.^37–40^ For example, in a community-based sample of Black adults (60% women), perceived stress was associated with a 37% greater risk of incident hypertension.^39^ Moreover, perceived racial discrimination across one’s lifetime was prospectively linked to incident hypertension.^37^ Several explanations are possible. First, neighborhood deprivation may act as a more stable and cumulative structural exposure, whereas psychosocial measures fluctuate over time and may be less predictive of long-term cardiovascular trajectories. Additionally, collinearity with socioeconomic covariates (e.g., education, insurance status) may have attenuated independent associations. Notably, one component of SV, women’s self-reported difficulties during pregnancy, remained associated with SBP, suggesting that certain subjective experiences of stress may still carry cardiovascular relevance for reproductive-aged women.

ADI and SV were not associated with incident hypertension over follow-up. These null findings may reflect limited accrual of new hypertension cases during the 2-7-year follow-up interval.^29,41^ BP elevations often precede the diagnostic threshold for hypertension, and postpartum BP represents a continuous phenotype influenced by both physiologic recovery from pregnancy and early environmental exposures.^42^ Yet, clinical hypertension represents a later, more multifactorial endpoint often requiring longer follow-up to emerge.^2,29^ Although the postpartum period is classically defined as the first year after pregnancy,^43^ longitudinal surveillance will be needed to capture early BP elevation prior to the onset of overt hypertension later in life.

Consistent with prior literature, women experienced APOs exhibited higher cardiometabolic risk in early pregnancy, including greater BMI, smoking prevalence, dyslipidemia, and BP than those without a history of APOs. APOs, particularly hypertensive disorders of pregnancy, may expose underlying vascular and metabolic vulnerability, and are associated with a twofold or greater risk of future hypertension and ischemic heart disease.^2,44^ Building on this evidence, our findings further demonstrate that neighborhood disadvantage contributes independently to postpartum BP elevations, even after accounting for APO history. These results suggest that adverse neighborhood contexts may interact with pregnancy-related biological vulnerability to shape long-term cardiometabolic trajectories.^45,46^

The potential pathways from neighborhood disadvantage to BP are likely multifactorial.^47^ Chronic stress related to economic hardship, crime exposure, and residential instability may promote neuroendocrine and inflammatory activation, and vascular dysfunction.^48^ Environmental conditions, including reduced access to green space, exposure to air pollution, and limited availability of healthy foods, further contribute to cardiometabolic risk.^49,50^ Structural barriers to healthcare access, such as provider shortages or transportation challenges, may also delay the early detection and management of elevated BP.^51,52^ Together, these pathways underscore that neighborhood disadvantage is a pervasive health context rather than simply a proxy of women’s individual behaviors.

### Clinical and Public Health Implications

Our approach allowed for assessment of how early exposures, both structural and individual, translate into measurable cardiovascular risk during the critical years after pregnancy, a time when many women transition away from intensive medical surveillance.^53^ Incorporating neighborhood-level risk indices such as ADI into postpartum cardiovascular risk assessment may enhance identification of women who could benefit from closer surveillance. Recent implementation work demonstrated the feasibility of integrating universal cardiovascular screening in the first year after delivery.^54^ Integration of CVD risk assessments in three large hospital networks in California resulted in screening 57-99% of eligible patients, greater rates than in a historical cohort.^55^ Given the structural barriers faced by women in disadvantaged neighborhoods, tailored follow-up and preventive care strategies (e.g., closer BP monitoring, earlier lifestyle counseling, and links to community resources) may be warranted.

At the public health and policy level, these results emphasize the need for interventions that extend beyond clinical care to address structural determinants of maternal cardiovascular health.^56,57^ Investments in housing stability, food access, and safe recreational spaces, could help reduce disparities in maternal CV risk. Because women from racially and socioeconomically marginalized groups are also disproportionately exposed to neighborhood disadvantage.^15,58^ Policies addressing residential segregation and other structural inequities may improve maternal cardiovascular health.^59,60^

Strengths of this study included a large, racially, and ethnically diverse cohort with prospective ascertainment of APOs, validated measures of psychosocial exposures, and standardized BP assessment. The longitudinal design enabled evaluation of pregnancy exposures in relation to BP measured years later. Still, several limitations must also be acknowledged. First, although the cohort was geographically diverse and racially and ethnically heterogeneous, participants were nulliparous women recruited at U.S. academic centers and may not fully represent multiparous women, individuals outside the United States, or those without access to early prenatal care. Thus, generalizability to broader populations should be considered with caution. Second, although we adjusted for key confounders, residual confounding by unmeasured factors (e.g., diet, physical activity, postpartum stressors) is possible. Third, the composite SV index incorporated both social exposures (e.g., discrimination, financial strain, social support) and psychological responses (e.g., perceived stress, anxiety, depressive symptoms). While this approach was intended to capture cumulative individual-level vulnerability, some psychological components may represent downstream consequences or mediators of social adversity rather than upstream exposures, potentially attenuating associations with postpartum BP. Fourth, SV was measured during pregnancy and may not fully capture more proximal exposures over follow-up, potentially attenuating associations. Fifth, BP was measured once postpartum, preventing evaluation of BP trajectories or variability. Sixth, attrition and missing data may introduce selection bias, although the final analytic sample remained large. Finally, the observational design precludes causal inference.

## Conclusions

In this prospective nuMoM2b-HHS cohort, area deprivation during pregnancy was associated with a small but durable influence on BP up to seven years after pregnancy. These results were independent of individual social and psychological vulnerabilities, which were not associated with BP at follow-up. These findings highlight the importance of structural environments in shaping early maternal cardiovascular trajectories and support the use of continuous BP measures as sensitive indicators of emerging risk. While replication of this investigation is needed, addressing the structural conditions that foster hypertension may be a central component of women’s cardiovascular health policy and practice. Future work separating upstream social exposures from downstream psychological responses may clarify whether individual-level vulnerability exerts effects through these pathways

## Data Availability

These data are available upon a reasonable request to the nuMoM2b investigators.

---

**Supplemental Table 1.** Baseline characteristics of participants by ADI quartiles.

|  | **Q1**<br>**(Least deprived)**<br>**N=934** | **Q2**<br>**N=960** | **Q3**<br>**N=934** | **Q4**<br>**(Most deprived)**<br>**N=881** | **p-value** |
| --- | --- | --- | --- | --- | --- |
| **Maternal age, years, mean (SD)** | 33.88 (4.96) | 31.47 (4.99) | 29.98 (5.22) | 27.74 (5.22) | <0.001 |
| **Age category, years, n (%)** |  |  |  |  | <0.001 |
| **18–21** | 5 (0.53) | 10 (1.04) | 29 (3.09) | 62 (6.96) |  |
| **22–35** | 491 (52.46) | 682 (70.89) | 720 (76.68) | 729 (81.82) |  |
| **>35** | 440 (47.01) | 270 (28.07) | 190 (20.23) | 100 (11.22) |  |
| **Race and ethnicity, n (%)** |  |  |  |  | <0.001 |
| **Non-Hispanic White** | 702 (75.00) | 766 (79.63) | 679 (72.31) | 289 (32.44) |  |
| **Non-Hispanic Black** | 30 (3.21) | 38 (3.95) | 90 (9.58) | 274 (30.75) |  |
| **Hispanic** | 109 (11.65) | 105 (10.91) | 104 (11.08) | 263 (29.52) |  |
| **Other** | 95 (10.15) | 53 (5.51) | 66 (7.03) | 65 (7.30) |  |
| **Relationship status: n(%)** |  |  |  |  | <0.001 |
| **Married, n (%)** | 786 (84.15) | 731 (76.15) | 594 (63.53) | 268 (30.21) |  |
| **Separated, divorced, or widowed** | 6 (0.64) | 7 (0.73) | 6 (0.64) | 8 (0.90) |  |
| **Single, never married** | 142 (15.20) | 222 (23.13) | 335 (35.83) | 611 (68.88) |  |
| **Education level** |  |  |  |  | <0.001 |
| **Less than high school graduate** | 6 (0.64) | 7 (0.73) | 29 (3.09) | 88 (9.88) |  |
| **High school graduate or GED** | 18 (1.92) | 62 (6.44) | 99 (10.54) | 193 (21.66) |  |
| **Some college credit, no degree** | 79 (8.44) | 159 (16.53) | 221 (23.54) | 224 (25.14) |  |
| **Associate/technical degree** | 316 (33.76) | 357 (37.11) | 256 (27.26) | 161 (18.07) |  |
| **Bachelor's degree** | 769 (82.16) | 624 (64.86) | 445 (47.39) | 251 (28.17) | — |
| **Graduate education** | 453 (48.40) | 267 (27.75) | 189 (20.13) | 90 (10.10) |  |
| **Income** |  |  |  |  |  |
| **<\$40,000** | 59 (6.88) | 134 (16.26) | 219 (29.16) | 336 (59.36) | |
| **\$40,000-\$74,999** | 134 (15.62) | 246 (29.85) | 251 (33.42) | 129 (22.79) | |
| **\$75,000-\$99,999** | 122 (14.22) | 152 (18.45) | 114 (15.18) | 48 (8.48) | |
| **\$100,000-\$149,999** | 218 (25.41) | 168 (20.39) | 114 (15.18) | 35 (6.18) | |
| **\$150,000-\$199,999** | 147 (17.13) | 78 (9.47) | 40 (5.33) | 14 (2.47) | |
| **\$200,000 or more** | 178 (20.75) | 46 (5.58) | 13 (1.73) | 4 (0.71) | |
| **Insurance, n (%)** |  |  |  |  | <0.001 |
| **Personal** | 244 (70.32) | 215 (61.78) | 116 (32.14) | 52 (9.00) |  |
| **Government** | 96 (27.67) | 120 (34.48) | 226 (62.60) | 500 (86.51) |  |
| **Military or Other** | 7 (2.02) | 13 (3.74) | 19 (5.26) | 26 (4.50) |  |
| **BMI, kg/m<sup>2</sup>, mean (SD)** | 24.68 (4.53) | 25.85 (5.57) | 26.92 (6.61) | 28.02 (7.27) | <0.001 |
| **Category: n (%)** |  |  |  |  | <0.001 |
| **<18** | 15 (1.66) | 13 (1.48) | 15 (1.83) | 22 (2.96) |  |
| **18 to 25** | 580 (64.16) | 516 (58.57) | 460 (56.03) | 349 (47.04) |  |
| **25 to <30** | 221 (24.45) | 249 (28.26) | 213 (25.94) | 237 (31.94) |  |
| **≥30** | 88 (9.73) | 103 (11.69) | 133 (16.20) | 134 (18.06) |  |
| **Visit 1 SBP, mmHg, mean (SD)** | 108.43 (9.46) | 109.28 (9.97) | 109.51 (10.45) | 108.42 (10.96) | 0.615 |
| **Visit 1 DBP, mmHg, mean (SD)** | 67.13 (7.69) | 67.54 (7.89) | 67.09 (8.47) | 66.38 (8.19) | 0.004 |
| **APOs: n (%)** | 131 (14.00) | 109 (11.33) | 146 (11.33) | 155 (17.40) | 0.002 |
| **Dyslipidemia: n (%)** | 150 (16.03) | 129 (13.41) | 135 (14.38) | 92 (10.33) | 0.006 |
| **Smoked during 3 months before pregnancy: n (%)** | 84 (8.97) | 113 (11.75) | 169 (18.00) | 206 (23.12) | <0.001 |
| **Family history of CVD: n (%)** | 568 (64.33) | 505 (55.25) | 520 (58.10) | 526 (64.94) | <0.001 |
| **Postpartum SBP, mmHg, mean (SD)** | 110.58 (10.71) | 110.77 (9.65) | 112.91 (10.58) | 112.67 (11.53) | <0.001 |
| **Postpartum DBP, mmHg, mean (SD)** | 71.10 (9.60) | 71.53 (9.10) | 73.59 (9.24) | 72.94 (9.91) | <0.001 |
| **Follow-up time, years, mean (SD)** | 3.20 (0.86) | 3.21 (0.88) | 3.12 (0.88) | 3.20 (0.94) | 0.485 |

**Notes:** Values are mean (SD) or n (%). p-values were from ANOVA or $\chi^2$ tests across quartiles.
Abbreviations: ADI = Area Deprivation Index; SBP = systolic blood pressure; DBP = diastolic blood pressure; Q1–Q4 = quartiles.

**Supplemental Table 2.** Baseline characteristics of participants by SV quartiles.

|  | **Q1**<br>**(Least SV)**<br>**N=791** | **Q2**<br>**N=792** | **Q3**<br>**N=791** | **Q4**<br>**(Most SV)**<br>**N=794** | **p-value** |
| --- | --- | --- | --- | --- | --- |
| **Maternal age, years, mean (SD)** | 31.49 (5.16) | 31.57 (5.13) | 31.00 (5.58) | 30.19 (5.93) | <0.001 |
| **Age category, years, n (%)** |  |  |  |  | <0.001 |
| **18–21** | 11 (1.39) | 8 (1.01) | 24 (3.03) | 31 (3.95) |  |
| **22–35** | 560 (70.71) | 549 (69.23) | 541 (68.31) | 554 (70.66) |  |
| **>35** | 221 (27.90) | 236 (29.76) | 227 (28.66) | 199 (25.38) |  |
| **Race and ethnicity, n (%)** |  |  |  |  | <0.001 |
| **Non-Hispanic White** | 581 (73.36) | 564 (71.12) | 547 (69.07) | 503 (64.16) |  |
| **Non-Hispanic Black** | 65 (8.21) | 64 (8.07) | 72 (9.09) | 102 (13.01) |  |
| **Hispanic** | 93 (11.74) | 100 (12.61) | 103 (13.01) | 129 (16.45) |  |
| **Other** | 53 (6.69) | 65 (8.20) | 70 (8.84) | 50 (6.38) |  |
| **Relationship status: n(%)** |  |  |  |  | <0.001 |
| **Married, n (%)** | 603 (76.33) | 590 (74.68) | 534 (67.68) | 395 (50.45) |  |
| **Separated, divorced, or widowed** | 4 (0.51) | 6 (0.76) | 9 (1.14) | 5 (0.64) |  |
| **Single, never married** | 183 (23.16) | 194 (24.56) | 246 (31.18) | 383 (48.91) |  |
| **Education level** |  |  |  |  | <0.001 |
| **Less than high school graduate** | 8 (1.01) | 22 (2.77) | 14 (1.77) | 41 (5.23) |  |
| **High school graduate or GED** | 51 (6.44) | 47 (5.93) | 63 (7.95) | 109 (13.90) |  |
| **Some college credit, no degree** | 121 (15.28) | 122 (15.38) | 160 (20.20) | 175 (22.32) |  |
| **Associate/technical degree** | 83 (10.48) | 85 (10.72) | 107 (13.51) | 103 (13.14) |  |
| **Bachelor's degree** | 264 (33.33) | 275 (34.68) | 224 (28.28) | 206 (26.28) |  |
| **Graduate education** | 264 (33.33) | 242 (30.52) | 224 (28.28) | 149 (19.01) |  |
| **Income** |  |  |  |  | <0.001 |
| **<\$40,000** | 119 (17.73) | 136 (19.94) | 143 (22.20) | 188 (31.60) | |
| **\$40,000-\$74,999** | 147 (21.91) | 179 (26.25) | 179 (27.80) | 153 (25.71) | |
| **\$75,000-\$99,999** | 113 (16.84) | 107 (15.69) | 92 (14.29) | 78 (13.11) | |
| **\$100,000-\$149,999** | 152 (22.65) | 130 (19.06) | 112 (17.39) | 97 (16.30) | |
| **\$150,000-\$199,999** | 68 (10.13) | 70 (10.26) | 59 (9.16) | 54 (9.08) | |
| **\$200,000 or more** | 72 (10.73) | 60 (8.80) | 59 (9.16) | 25 (4.20) | |
| **Insurance, n (%)** |  |  |  |  |  |
| **Personal** | 140 (51.28) | 151 (47.94) | 141 (45.05) | 127 (31.20) |  |
| **Government** | 123 (45.05) | 147 (46.67) | 158 (50.48) | 267 (65.60) |  |
| **Military or Other** | 10 (3.66) | 17 (5.40) | 14 (4.47) | 13 (3.19) |  |
| **BMI, kg/m<sup>2</sup>, mean (SD)** | 25.55 (5.62) | 25.93 (6.02) | 26.13 (6.08) | 27.22 (6.73) | 0.009 |
| **Category: n (%)** |  |  |  |  | 0.010 |
| **<18** | 15 (2.04) | 14 (1.93) | 18 (2.54) | 13 (1.90) |  |
| **18 to 25** | 449 (61.17) | 439 (60.39) | 405 (57.20) | 355 (51.98) |  |
| **25 to <30** | 195 (26.57) | 184 (25.31) | 194 (27.40) | 200 (29.28) |  |
| **≥30** | 75 (10.22) | 90 (12.38) | 91 (12.85) | 115 (16.84) |  |
| **Visit 1 SBP, mmHg, mean (SD)** | 108.41 (9.93) | 109.17 (9.89) | 109.31 (10.48) | 108.78 (10.55) | 0.286 |
| **Visit 1 DBP, mmHg, mean (SD)** | 66.68 (7.90) | 67.03 (7.80) | 67.08 (8.20) | 67.29 (8.32) | 0.521 |
| **APOs: n (%)** | 98 (12.37) | 85 (10.72) | 118 (14.90) | 134 (17.09) | 0.001 |
| **Dyslipidemia: n (%)** | 104 (13.13) | 112 (14.12) | 106 (13.38) | 112 (14.29) | 0.860 |
| **Smoked during 3 months before pregnancy: n (%)** | 81 (10.23) | 89 (11.22) | 119 (15.03) | 189 (24.11) | <0.001 |
| **Family history of CVD: n (%)** | 473 (62.07) | 462 (61.19) | 461 (62.81) | 420 (57.93) | 0.235 |
| **Postpartum SBP, mmHg, mean (SD)** | 111.57 (10.33) | 111.31 (9.96) | 111.32 (11.21) | 111.44 (10.46) | <0.001 |
| **Postpartum DBP, mmHg, mean (SD)** | 71.48 (8.99) | 72.15 (9.17) | 71.94 (10.04) | 72.55 (9.42) | <0.001 |
| **Follow-up time, years, mean (SD)** | 3.14 (0.87) | 3.17 (0.87) | 3.14 (0.87) | 3.17 (0.90) | 0.785 |

**Notes:** Values are mean (SD) or n (%). p-values were from ANOVA or $\chi^2$ tests across quartiles.
Abbreviations: ADI = Area Deprivation Index; SBP = systolic blood pressure; DBP = diastolic blood pressure; Q1-Q4 = quartiles.

**Supplemental Table 3.**
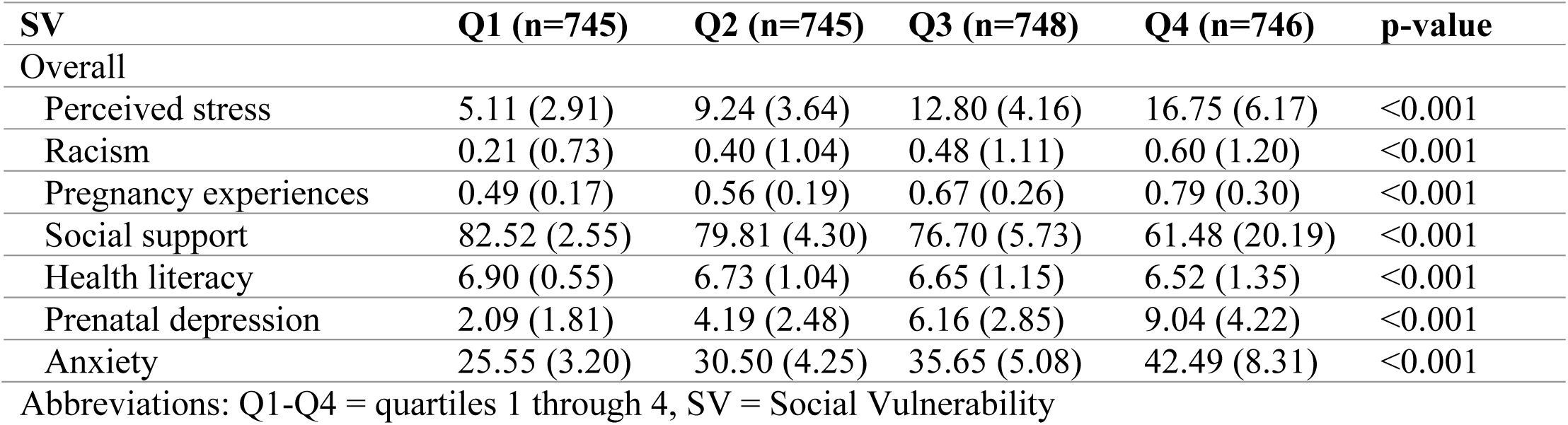
SV quartiles by component.

**Supplemental Table 4.**
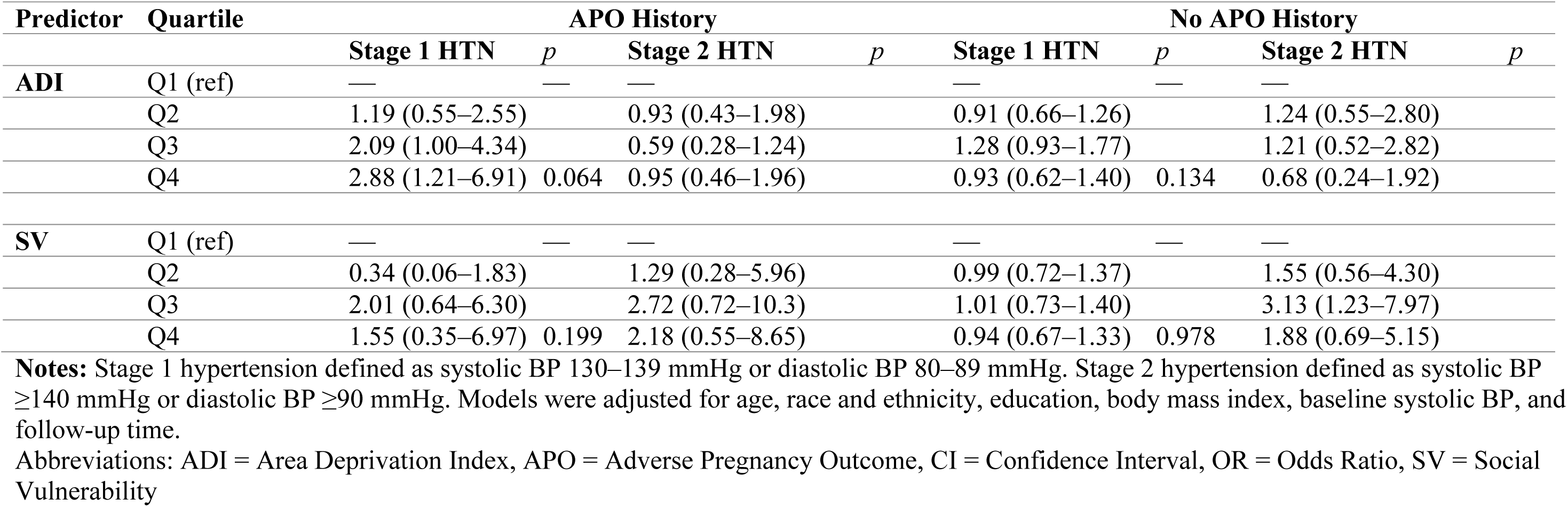
Associations of ADI and SV Quartiles with Incident Hypertension at 2–7 Years Postpartum, Overall and by APO History.

**Supplemental Table 5.** Associations of SV and ADI with blood pressure at 2-7 years postpartum, by APO history.

| APO |  |  |  |  |  |  | No APO |  |  |  |  |  |
| --- | --- | --- | --- | --- | --- | --- | --- | --- | --- | --- | --- | --- |
| Separate ADI and SV models |  |  |  |  |  |  |  |  |  |  |  |  |
|  | SBP |  |  | DBP |  |  |  | SBP |  |  | DBP |  |
|  | B (SE) | 95% CI | p | B (SE) | 95% CI | p | B (SE) | 95% CI | p | B (SE) | 95% CI | p |
| ADI | 0.05 (0.02) | 0.01,0.10 | 0.015 | 0.04 (0.02) | 0.01,0.08 | 0.039 | 0.03 (0.01) | 0.01,0.04 | <0.001 | 0.02 (0.01) | 0.01,0.04 | 0.003 |
| SV | -0.03 (0.55) | -1.11,1.06 | 0.961 | -0.44 (0.49) | -1.41,0.52 | 0.367 | -0.45 (0.20) | -0.84,-0.06 | 0.023 | -0.30 (0.17) | -0.64,0.05 | 0.089 |
| ADI and SV models |  |  |  |  |  |  |  |  |  |  |  |  |
|  | SBP |  |  | DBP |  |  |  | SBP |  |  | DBP |  |
|  | B (SE) | 95% CI | p | B (SE) | 95% CI | p | B (SE) | 95% CI | p | B (SE) | 95% CI | p |
| ADI | 1.62 (0.66) | 0.32,2.92 | 0.015 | 1.21 (0.59) | 0.05,2.38 | 0.042 | 0.80 (0.21) | 0.38,1.23 | <0.001 | 0.55 (0.19) | 0.18,0.92 | 0.004 |
| SV | 0.01 (0.66) | -1.08,1.09 | 0.997 | -0.36 (0.50) | -1.34,0.61 | 0.465 | -0.42 (0.20) | -0.81,-0.02 | 0.038 | -0.26 (0.18) | -0.61,0.09 | 0.144 |

**Notes:**
P-values are based on covariate-adjusted linear mixed models of ADI, SV, or the ADI and SV composite.
Covariates included age, race and ethnicity, education, body mass index, baseline systolic blood pressure, and follow-up time.
Abbreviations: ADI, Area Deprivation Index; APO, adverse pregnancy outcome; DBP, diastolic blood pressure; SBP, systolic blood pressure; SV, social vulnerability.

**Supplemental Table 6.**
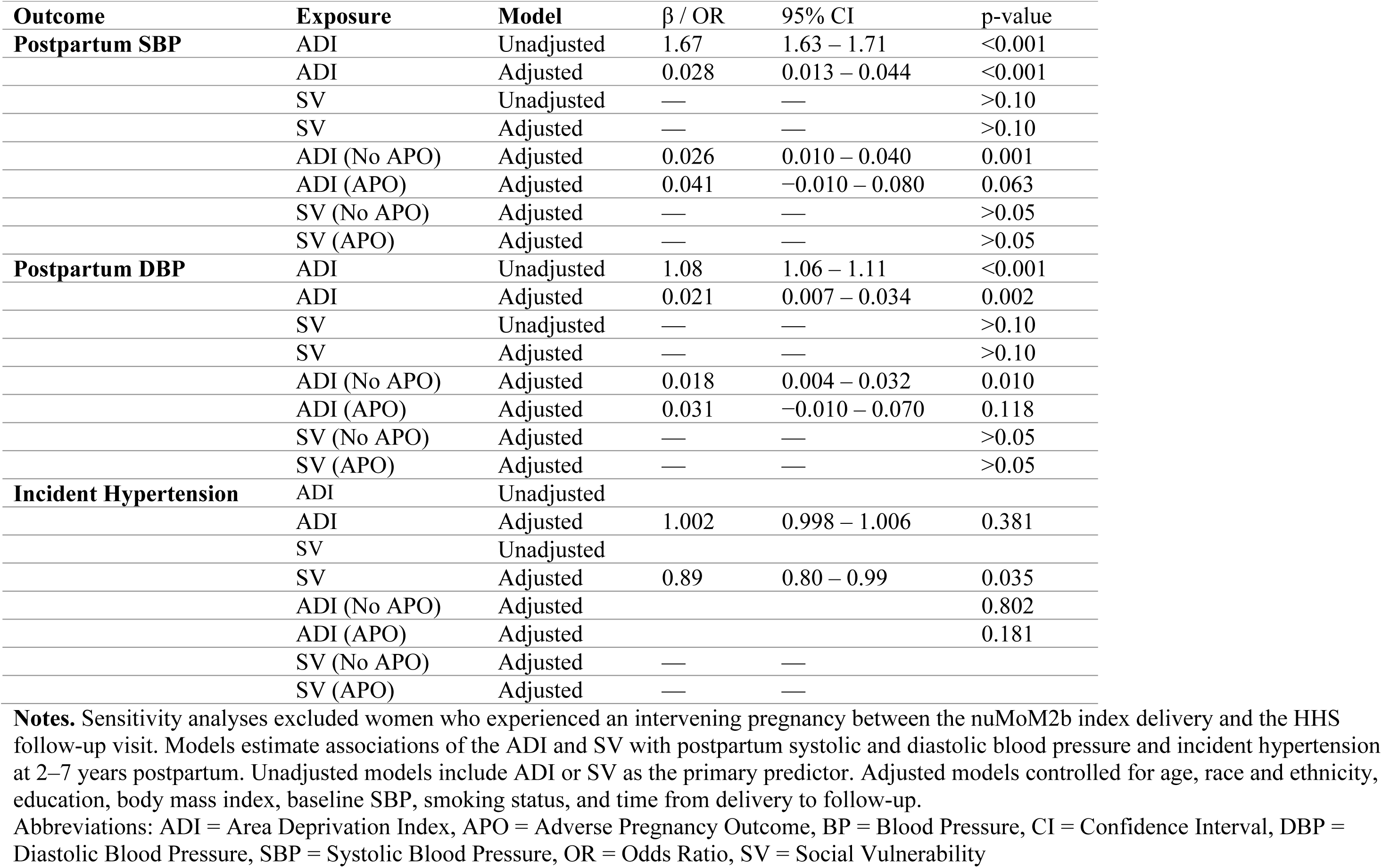
Sensitivity Analyses Excluding Women with an Intervening Pregnancy.

| Outcome | Exposure | Model | $\beta$ / OR | 95% CI | p-value |
| --- | --- | --- | --- | --- | --- |
| **Postpartum SBP** | ADI | Unadjusted | 1.67 | 1.63 – 1.71 | <0.001 |
|  | ADI | Adjusted | 0.028 | 0.013 – 0.044 | <0.001 |
|  | SV | Unadjusted | — | — | >0.10 |
|  | SV | Adjusted | — | — | >0.10 |
|  | ADI (No APO) | Adjusted | 0.026 | 0.010 – 0.040 | 0.001 |
|  | ADI (APO) | Adjusted | 0.041 | –0.010 – 0.080 | 0.063 |
|  | SV (No APO) | Adjusted | — | — | >0.05 |
|  | SV (APO) | Adjusted | — | — | >0.05 |
| **Postpartum DBP** | ADI | Unadjusted | 1.08 | 1.06 – 1.11 | <0.001 |
|  | ADI | Adjusted | 0.021 | 0.007 – 0.034 | 0.002 |
|  | SV | Unadjusted | — | — | >0.10 |
|  | SV | Adjusted | — | — | >0.10 |
|  | ADI (No APO) | Adjusted | 0.018 | 0.004 – 0.032 | 0.010 |
|  | ADI (APO) | Adjusted | 0.031 | –0.010 – 0.070 | 0.118 |
|  | SV (No APO) | Adjusted | — | — | >0.05 |
|  | SV (APO) | Adjusted | — | — | >0.05 |
| **Incident Hypertension** | ADI | Unadjusted |  |  |  |
|  | ADI | Adjusted | 1.002 | 0.998 – 1.006 | 0.381 |
|  | SV | Unadjusted |  |  |  |
|  | SV | Adjusted | 0.89 | 0.80 – 0.99 | 0.035 |
|  | ADI (No APO) | Adjusted |  |  | 0.802 |
|  | ADI (APO) | Adjusted |  |  | 0.181 |
|  | SV (No APO) | Adjusted | — | — |  |
|  | SV (APO) | Adjusted | — | — |  |

**Notes.** Sensitivity analyses excluded women who experienced an intervening pregnancy between the nuMoM2b index delivery and the HHS follow-up visit. Models estimate associations of the ADI and SV with postpartum systolic and diastolic blood pressure and incident hypertension at 2–7 years postpartum. Unadjusted models include ADI or SV as the primary predictor. Adjusted models controlled for age, race and ethnicity, education, body mass index, baseline SBP, smoking status, and time from delivery to follow-up.
Abbreviations: ADI = Area Deprivation Index, APO = Adverse Pregnancy Outcome, BP = Blood Pressure, CI = Confidence Interval, DBP = Diastolic Blood Pressure, SBP = Systolic Blood Pressure, OR = Odds Ratio, SV = Social Vulnerability

## Acknowledgements

We would like to thank all the nuMoM2b participants for their time.

